# What the COVID-19 pandemic teaches us about modelling epidemics: Percolation versus SEIR

**DOI:** 10.1101/2025.11.13.25340147

**Authors:** Jean-François Mathiot, Laurent Gerbaud, Vincent Breton

## Abstract

From the study of four typical periods of the SARS-CoV-2 virus propagation in France in 2019, 2020 and 2021, we investigate the role played by the local spread of the epidemic as compared to a long-range propagation. While the local spread of the epidemic is driven by the social relationships in the population under consideration, the long-range propagation does not involve any close connection - either geographical or sociological - between individuals. This comparison is done using the PERCOVID model based on percolation theory as compared to the SEIR model based on the global evolution of the number of persons in various compartments. This study is a first step in using PERCOVID as a new tool in public health for the early detection of respiratory track epidemics in countries with very different social behaviors.

## Introduction

The modelling of how epidemics propagate is essential for identifying the best strategies to control their diffusion in a given population. In this respect, the COVID-19 pandemic has provided a tremendous testing ground for the various models already available in the literature. As compared to the annual flu epidemics, it has provided almost all possible situations which a viral infection can encounter: initial underground propagation during several months in a non-vaccinated population, exponential explosion and influence of drastic mobility restrictions during lockdown periods, propagation during summer vacations with a very large mobility of the population, propagation of variants with increasing virulence, relative effectiveness of immunization, loss of natural or vaccination immunity with time, etc. These different situations must of course be accounted for by epidemic modelling. The model must integrate the intrinsic epidemiological properties of the virus, and in particular its virulence. It also has to take into account the way the virus propagates, either through respiratory track, by contact or by long distance contamination. Knowledge of these different key points is required to construct a reliable epidemiological modelling.

The most widely used methods to predict COVID-19 pandemic are compartmental models [1]. In these models, the population is separated into mutually exclusive groups, or compartments, based on their disease status [2]. To this family belong the so-called SEIR (Susceptible, Exposed, Infected and Recovered) models. Percolation models focus on the step-by-step propagation of a given event in a medium made up of a very large number of elements [3]. Quite naturally, these models have been applied to the step-by-step spread of an epidemic in a given population, through the formation of clusters [4].

Most researchers adopted the SEIR model and its variants to forecast COVID-19 metrics due to its simplicity, its prevalence among academics and the long-standing experience of epidemiologists in using them [5,6]. However, limitations of their performances, as well as biases and tendencies to over forecast have been already documented [7–10]. We compare in this study the prediction of SEIR models with a recently proposed percolation type model [11], called PERCOVID during different periods of the SARS-CoV-2 virus propagation in France in 2019, 2020 and 2021.

## Methods

The PERCOVID simulation is based on a stochastic model defined on a discretized *D*-dimensional network while the SEIR models are deterministic models of the time evolution of continuous functions. If all other hypotheses are the same, this difference in the numerical treatment does not however lead to significant differences in the numerical results provided the number of individuals considered in the model is large enough. This is usually the case when we consider the propagation of an epidemic on a large territory. The way the two models differ can be easily understood if we consider, on the one hand, the definition of the initial state, and on the other hand the propagation patterns of the virus. In a precedent work, we checked that PERCOVID was a good predictor of the dynamics of the COVID19 epidemics [11].

In SEIR models, one should first fix the number of individuals in each compartment, that is the number of Susceptible, Exposed, Infected and Recovered persons. The dynamics of the propagation is thus determined by the reproduction rate, denoted by R, which defines the mean number of contaminations an infected person can induce during the time of its infection. The reproduction rate is thus a global parameter which encompasses parameters of very different nature, like the virulence of the virus or the sociological behavior of the population.

In the PERCOVID model, we should also define the initial number of persons of each type. However, PERCOVID is able to handle the very early stage of an epidemic, as we can only choose an initial number of infected persons sufficient to start a long-time propagation of the virus [2,11]. The dynamics of the epidemic is then determined by two sets of parameters. The first set refers to the virulence of the virus, an intrinsic property independent of any sociological considerations. It is called intrinsic infectiousness in PERCOVID, and denoted by *r*. The second one characterizes the social behavior of the population on the territory in which the virus propagates. It is mainly determined by the density, intensity and variety of social relationships [11].

Once the initial state is defined, the two models differ drastically in the way the virus propagation is computed. By construction in SEIR models, the infection of a susceptible person by an infected one is completely independent of any possible relationship between these two persons, whether geographical or sociological. On the contrary, PERCOVID, as a percolation model, relies entirely on the social relationships each person can have on the territory under consideration. The intensity of these relationships is the highest for individuals living in the same household, then decreases for closest neighbors and decreases further for next-to-closest neighbors [11]. Note that in PERCOVID the concept of neighbor is not geographical but reflects a significant social relationship between two individuals belonging to different households.

From the above considerations, the comparison between PERCOVID and SEIR models proceeds as follows. We have identified four ten-week crucial periods between December 2019 and November 2021 during the COVID-19 epidemic in France. These periods correspond to different virus strains and different intensities of social relationships due to public health non-pharmaceutical interventions (such as complete or partial lockdowns). Pseudo-data for these four periods were generated from the complete PERCOVID simulation of the epidemics in France [11]. In this simulation, the weekly number of newly hospitalized persons in France (the most reliable data available all along the epidemic) is used to fix the intrinsic infectiousness *r* of the initial strain and the successive variants. The simulation predicts the initial number of susceptible, exposed, infected and recovered persons (together with the number of vaccinated persons, if any) at the beginning of each 10-week period.

The SEIR-LIKE model aims at describing the epidemic on the same network and with the same number of daily relationships as PERCOVID, but with a different infection rule. In the SEIR-LIKE simulation, a person can infect randomly a susceptible one everywhere on the network, and not only in its close neighborhood as PERCOVID model does. This mimics the fundamental difference between PERCOVID and SEIR models, all other considerations being the same. In order to settle the initial conditions for the SEIR-LIKE simulation, we fix the corresponding intrinsic infectiousness of this model, denoted by *r*_*seir*_, to get the same slope in both models for the daily number of newly infected persons during the first two weeks of each period. This corresponds typically to the method used for the determination of the reproduction rate R for any SEIR simulation [12].

As we shall see below, the value of the intrinsic infectiousness in both models, *r* and *r*_*seir*_, can be quite different. This difference may induce a difference in the number of infected persons at the beginning of each period. In order to fix the same initial number of infected persons in the PERCOVID and SEIR-LIKE simulations, we therefore renormalize the number of infected persons in the SEIR-LIKE simulation to get the same absolute number as in the PERCOVID one during the first two-weeks of each period. Once this initial state is defined, together with the intrinsic infectiousness for each simulation, we can thus compare the prediction of both simulations for the remaining eight weeks of each period.

## Results

### Determination of initial conditions for the SEIR-LIKE simulation

The daily number of newly infected persons for the first two weeks of the four periods considered in this work is shown on figure 1 for both PERCOVID and SEIR-LIKE simulations. The intrinsic infectiousness in the SEIR-LIKE simulation together with the normalization factor needed to get the same number of infected persons during these initial periods are reported in table 1. In the SEIR-LIKE simulation, a contagious person can infect any susceptible person in the network while contagion is limited to the household and social neighbors in the PERCOVID model. As a consequence, to get the same slope for the daily number of newly infected persons, the virus virulence r_seir_ is always lower than its virulence in the PERCOVID model. The results correspond to the mean value of 100 simulations.

**Table 1.** Intrinsic infectiousness for the initial strain and the *δ* variant, together with the renormalization factor for each period in the SEIR-LIKE simulations as compared to the PERCOVID one (last column). We also indicate the corrections, as a percentage, to the intrinsic infectiousness in the PERCOVID simulation

| Period | 1 | 2 | 3 | 4 | PERCOVID |
| --- | --- | --- | --- | --- | --- |
| Initial strain | 0.053<br>62% | 0.031<br>78% | 0.095<br>32% | - | 0.14 |
| $\delta$ variant | - | - | - | 0.30<br>29% | 0.42 |
| Renormalization | 1.48 | 1.34 | 1.06 | 1.02 | - |

**Figure 1.**
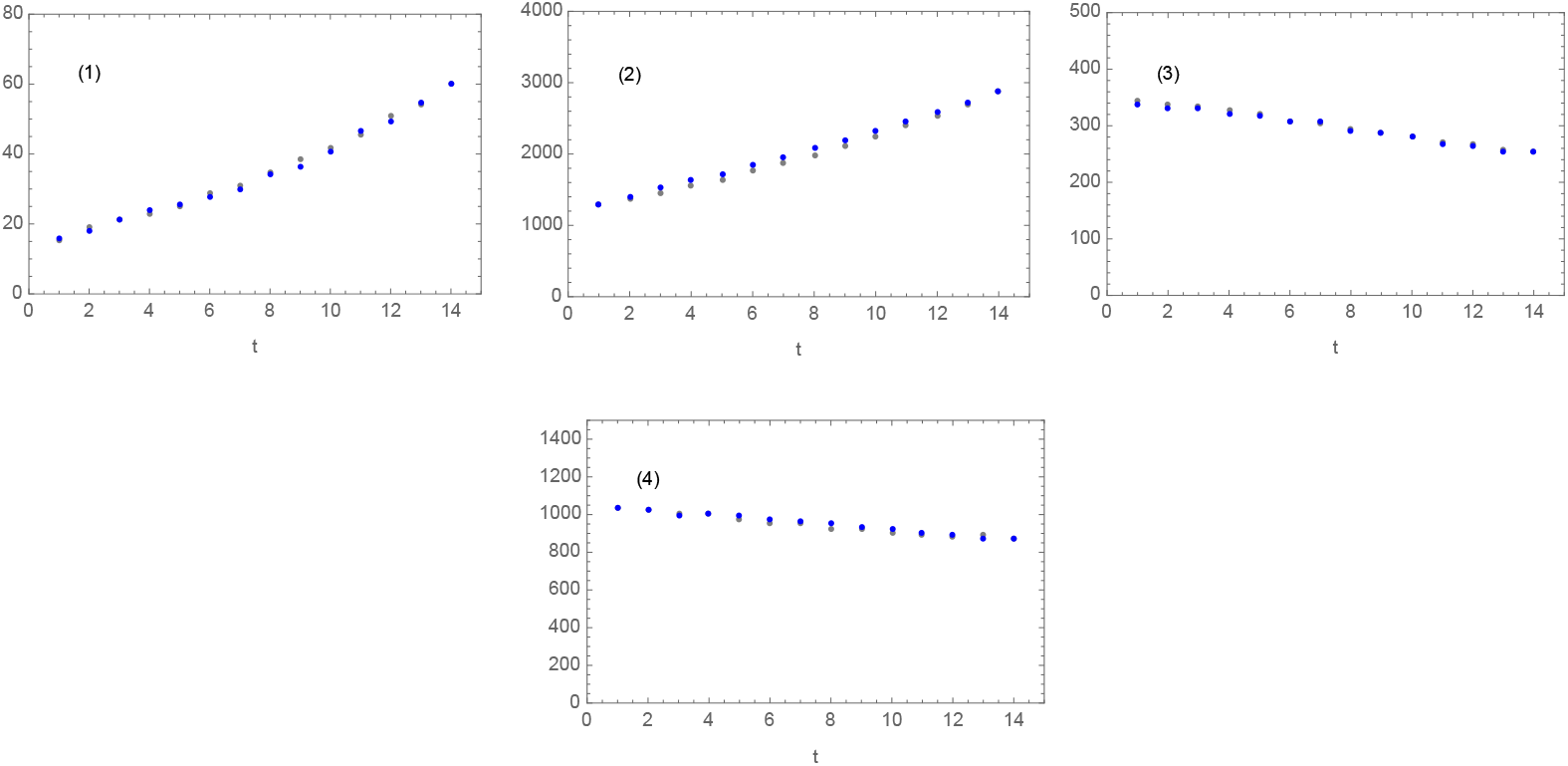
Comparison of the daily number of newly infected persons as a function of time (expressed in days) between PERCOVID (gray points) and SEIR-LIKE (blue points) simulations for the first two weeks of the four periods studied (labelled 1 to 4). These periods are detailed below.

### Period 1: December 9, 2019 / February 16, 2020

This period corresponds to the initial underground propagation of the virus and starts several weeks before the first official case was reported in France, the 24th of January 2020. We show on figure 2 the comparison between PERCOVID and SEIR-LIKE simulations for the reproduction number R, the symptomatic weekly incidence rate and the percentage of secondary infections. This period corresponds to the propagation of the virus in an almost non-protected population, with no available vaccination, no natural immunity, and none of the non-pharmaceutical interventions enforced such as the obligation of wearing a mask.

**Figure 2.**
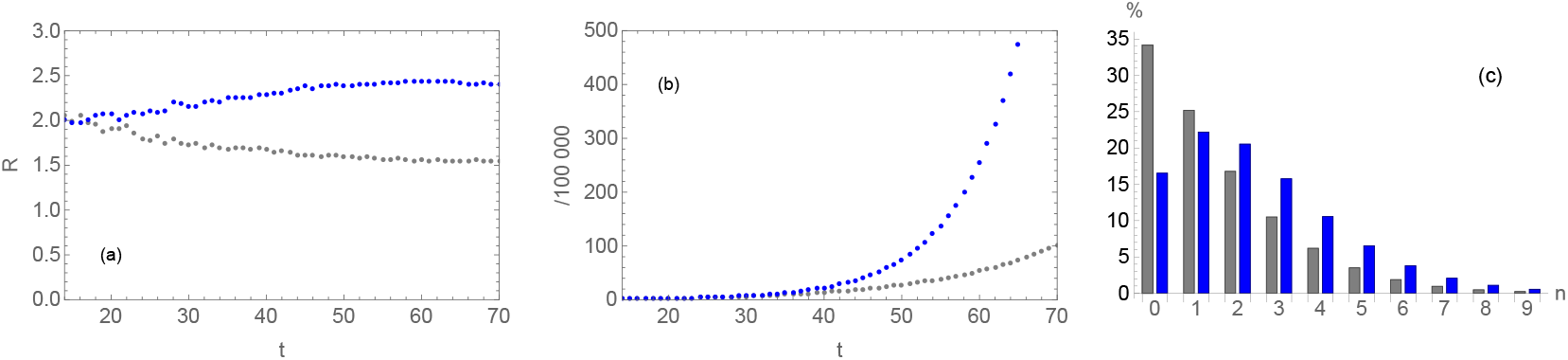
Comparison between PERCOVID (gray) and SEIR-LIKE (blue) simulations for the period 1, for the reproduction number R (a), the symptomatic weekly incidence rate (b) and the percentage of secondary infections (c). Time t is expressed in days.

- The intrinsic infectiousness in the SEIR-LIKE simulation, as compared to PERCOVID, is reduced by 62%, with a normalization factor of 1.48, as given in table 1. This large difference highlights the influence of the different propagation pattern in both simulations.
- The overall dynamics of the epidemic is governed by the reproduction rate R. It is large - about 2 - at the beginning of the period and shows a quite different pattern for PERCOVID and SEIR-LIKE simulations. SEIR-LIKE predicts a regular increase of the reproduction rate while PERCOVID predicts a slow but regular decrease. This decrease is due to the decreasing number of non-infected individuals in the local (social) environment of each infected person in the absence of a large mobility of persons. On the contrary, the maximum of R at the end of the period in the SEIR-LIKE simulation is due to the extremely rapid contamination of susceptible persons all over the territory.
- Despite the large reduction of the intrinsic infectiousness in the SEIR-LIKE simulation, there is an extremely rapid growth of the epidemic in this case, very different from the PERCOVID predicted behavior. This is just a consequence of the different predictions for the reproduction number R, as mentioned above. This leads to a very large overestimate of the number of infected persons at the end of this period in the SEIR-LIKE simulation.
- As a direct consequence, the SEIR-LIKE model predicts a very different pattern for the distribution of secondary infections than PERCOVID. Due to the long-distance contamination in the SEIR-LIKE simulation, the average number of secondary infections is much larger in this case, as indicated on figure 2.c. While about 35% of infected persons do not infect anybody according to the PERCOVID simulation, this rate is about only 15% in the SEIR-LIKE simulation. This is indeed in perfect agreement with the extremely rapid propagation of the virus in this case.

### Period 2: February 3, 2020/ April 12, 2020

This period corresponds to the very large, exponential, expansion of the virus once it has spread all over the territory during the first period of underground propagation. Results are shown on figure 3. The first lockdown in France occurs just at time t=44 days after the beginning of this period.

**Figure 3.**
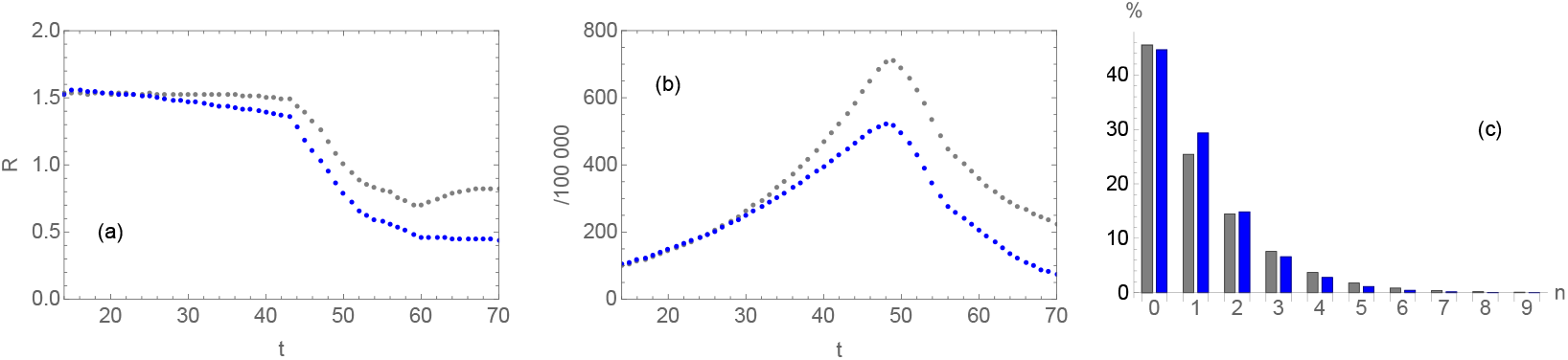
Comparison between PERCOVID (gray) and SEIR-LIKE (blue) simulations for the period 2, for the reproduction number R (a), the symptomatic weekly incidence rate (b) and the percentage of secondary infections (c). Time t is expressed in days.

- The intrinsic infectiousness in the SEIR-LIKE simulation is reduced by 78%, with a normalization factor of 1.34, as compared to PERCOVID (table 1). This behavior is similar to what has been found in period 1. This corresponds indeed to a somewhat similar situation, with however a regular spread of the epidemic over the whole territory after more than two months of virus propagation. This large spread implies a slightly different dynamics as compared to period 1.
- The reproduction number R is very similar in both simulations. This is a direct consequence of the large spread of the epidemic over the whole territory, making the difference between the propagation pattern of both simulations less stringent. This also makes the propagation patterns of both simulations more similar. Since the intrinsic infectiousness in the SEIR-LIKE simulation is much reduced as compared to the PERCOVID one, the incidence rate in the population is thus progressively reduced in the SEIR-LIKE simulation. It thus overestimates the impact of the confinement by about 30% at the end of this period.
- The percentage of secondary infections is very similar in both simulations. This is a direct consequence of the large dissemination of the virus all over the territory which implies a similar environment in both simulations.

### Period 3: June 15, 2020 / August 23, 2020

This period corresponds to summer vacations after the first epidemic wave, with second underground epidemic slowly progressing, without major outbreaks, so preparing a second epidemic wave on a much larger geographic basis. Figure 4 displays comparison of SEIR-LIKE and PERCOVID predictions during 2020 summer vacations, with a very large mobility of persons and hence a very large dissemination of infected persons. This should lead to a dynamic similar to what we discussed above in period 2, with however a less pronounced effect since the number of infected persons is much reduced after the first lockdown, as can be seen by comparing the scales of figures 3.b and 4.b. These features translate in a smaller reduction factor of the intrinsic infectiousness in the SEIR-LIKE simulation as compared to the previous periods. It is 32%, with a renormalization factor of 1.06 (see table 1).

**Figure 4.**
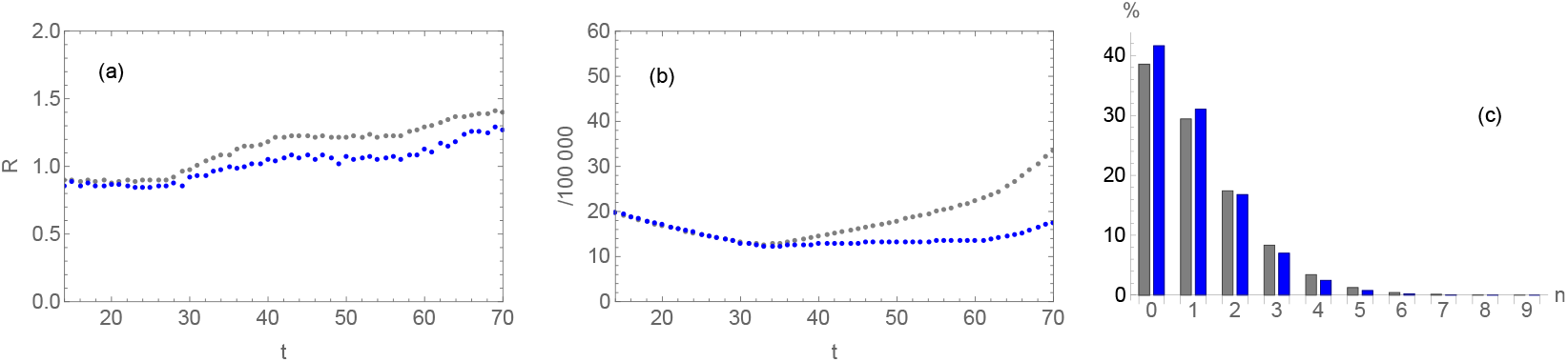
Comparison between PERCOVID (gray) and SEIR-LIKE (blue) simulations for the period 3, for the reproduction number R (a), the symptomatic weekly incidence rate (b) and the percentage of secondary infections (c). Time t is expressed in days.

- The reproduction number R is also almost the same in both simulations, with however a very slight decrease in the SEIR-LIKE simulation as compared to the PERCOVID one in the second part of the period. This period corresponds to an increase of the density of social relationships during summer which thus translates in a slightly larger value for R in the PERCOVID simulation since the intrinsic infectiousness is larger in this simulation than in the SEIR-LIKE one.
- The reduction of R in the SEIR-LIKE simulation induces a smaller infection pattern at the end of this period. This leads to an underestimate of the symptomatic weekly infection rate by about 40% at the end of this period
- The percentage of secondary infections is also almost identical in both simulations, as expected from the dynamic of the epidemic during this period.

### Period 4: September 13, 2021 / November 21, 2021

This last period corresponds to the rapid spread of the delta variant, with a very high intrinsic infectiousness, together with the progressive loss of vaccine efficiency and immunity. We thus recover the conditions found in the first period of propagation of the virus, as far as the delta variant is concerned. Results are shown on figure 5.

**Figure 5.**
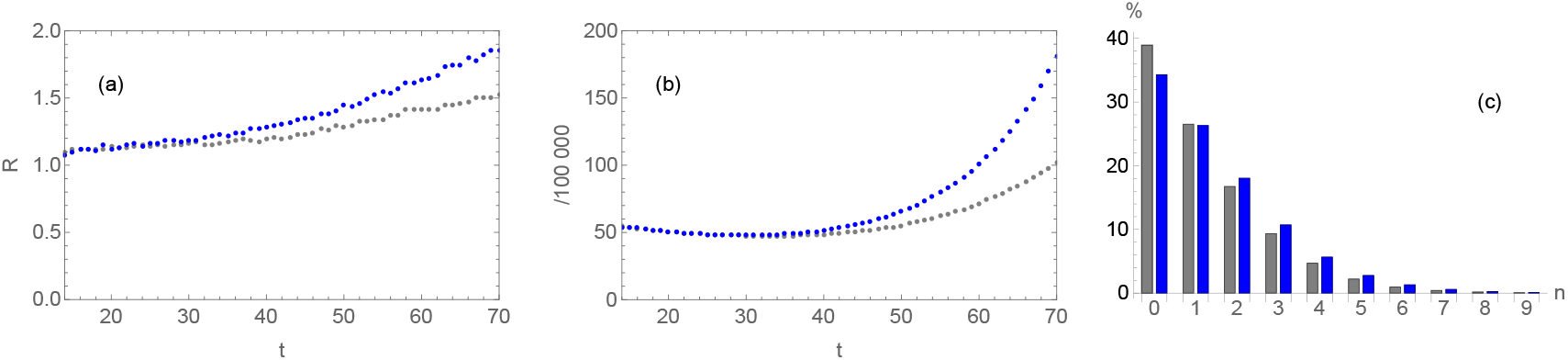
Comparison between PERCOVID (gray) and SEIR-LIKE (blue) simulations for the period 4, for the reproduction number R (a), the symptomatic weekly incidence rate (b) and the percentage of secondary infections (c). Time t is expressed in days.

- The intrinsic infectiousness is only slightly reduced in the SEIR-LIKE simulation as compared to the PERCOVID one. It is 29% smaller with a renormalization factor of about 1.
- There is a regular increase of the reproduction number R due to the large intrinsic infectiousness of the delta variant. This increase is however much larger in the SEIR-LIKE simulation, just as in the first period, with thus a much faster increase of the symptomatic weekly incidence rate. This leads also to a large overestimate - about 80% - of the propagation of the virus at the end of this period. The propagation of the delta variant, with its large intrinsic infectiousness together with a less efficient vaccine, just looks like the propagation of the initial strain in the first period. This overestimate is however much less pronounced since there is already a large natural and vaccinal immunity together with a very large propagation of the virus all over the territory as compared to the very beginning of the epidemic.
- This similarity with the dynamics discussed in period 1 translates also in the distribution of secondary infections, with a sizeable increase of the average number of secondary infections, and a subsequent decrease of the number of persons with zero secondary infections in the SEIR-LIKE simulation.

### Influence of initial conditions

The results discussed above were obtained for an initial state in the PERCOVID simulation with only 14 infected persons out of about 5.6 million individuals that represent a tenth of the French population. This corresponds to an incidence rate (number of infected persons per 100.000 habitants) T_0_ very close to 0.25. In order to investigate the influence of these initial conditions on the comparison between PERCOVID and SEIR-LIKE simulations, the predictions of both simulations were compared with two different initial conditions: a larger rate of initial infections (multiplication by a factor of ten), and a non-zero rate of initial vaccination, with a coverage of 25% and 50% of vaccinated persons in the population. We focus for this comparison on the first period corresponding to the very beginning of the epidemic propagation.

We show in table 2 the reduction of the intrinsic infectiousness in the SEIR-LIKE simulations, together with the corresponding renormalization factor to get the same initial number of infected persons as compared to the PERCOVID simulation during the first two-weeks of the first period. The 10-fold increase in the initial number of infected persons does not impact the comparison between the two simulations, with identical intrinsic infectiousness and renormalization factor in both cases, as shown in the first two columns of table 2. The difference between the two simulations on the other hand is slightly reduced when the number of vaccinated persons increases, as shown in the last three columns of table 2. Figure 6 shows the daily number of newly infected persons in both simulations during the initial two weeks of the period. We checked again that the incidence rates are almost identical in both simulations once the intrinsic infectiousness and renormalization factors in the SEIR-LIKE simulation are adjusted. This fixes the initial conditions for the comparison between these two simulations for the remaining eight weeks of this period.

**Table 2.** Intrinsic infectiousness and renormalization factor for three different vaccination rates (0%, 25%, 50%) in the PERCOVID and SEIR-LIKE simulations and for two different initial incidence rate (number of infected persons per 100.000 of the population) T_0_=0.25 and T_0_=2.5.

| | 0% ($T_0=0.25$) | 0% ($T_0=2.5$) | 25% ($T_0=2.5$) | 50% ($T_0=2.5$) |
| --- | --- | --- | --- | --- |
| SEIR | 0.053<br>62% | 0.053<br>62% | 0.060<br>57% | 0.068<br>51% |
| Renormalization | 1.48 | 1.48 | 1.38 | 1.20 |

**Figure 6.**
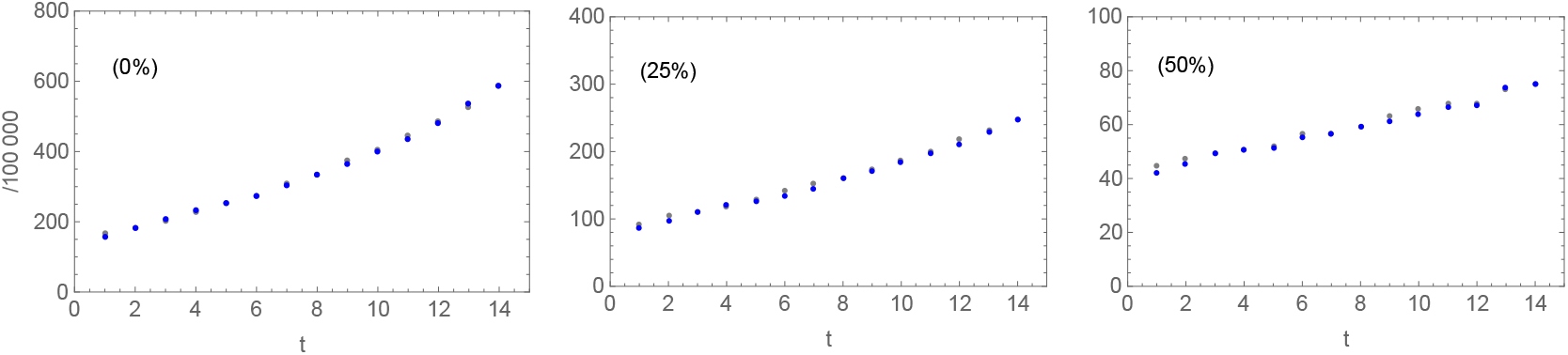
Comparison between the daily number of newly infected persons during the initial two weeks calculated in the PERCOVID (gray points) and SEIR-LIKE (blue points) simulations for three different vaccination rates (0%, 25%, 50%) and for an initial incidence rate of T=2.5.

In order to understand more precisely the effect of the vaccination rate on the way the virus propagates in both PERCOVID and SEIR-LIKE simulations, we show on figures 7 and 8 the same observables as considered above but with a rate of initial vaccination of 0%, 25% and 50% and an initial incidence rate of T_0_=2.5. The patterns we can see from these figures are in line with what has been seen in the first period shown on figure 2, with however few differences:

**Figure 7.**
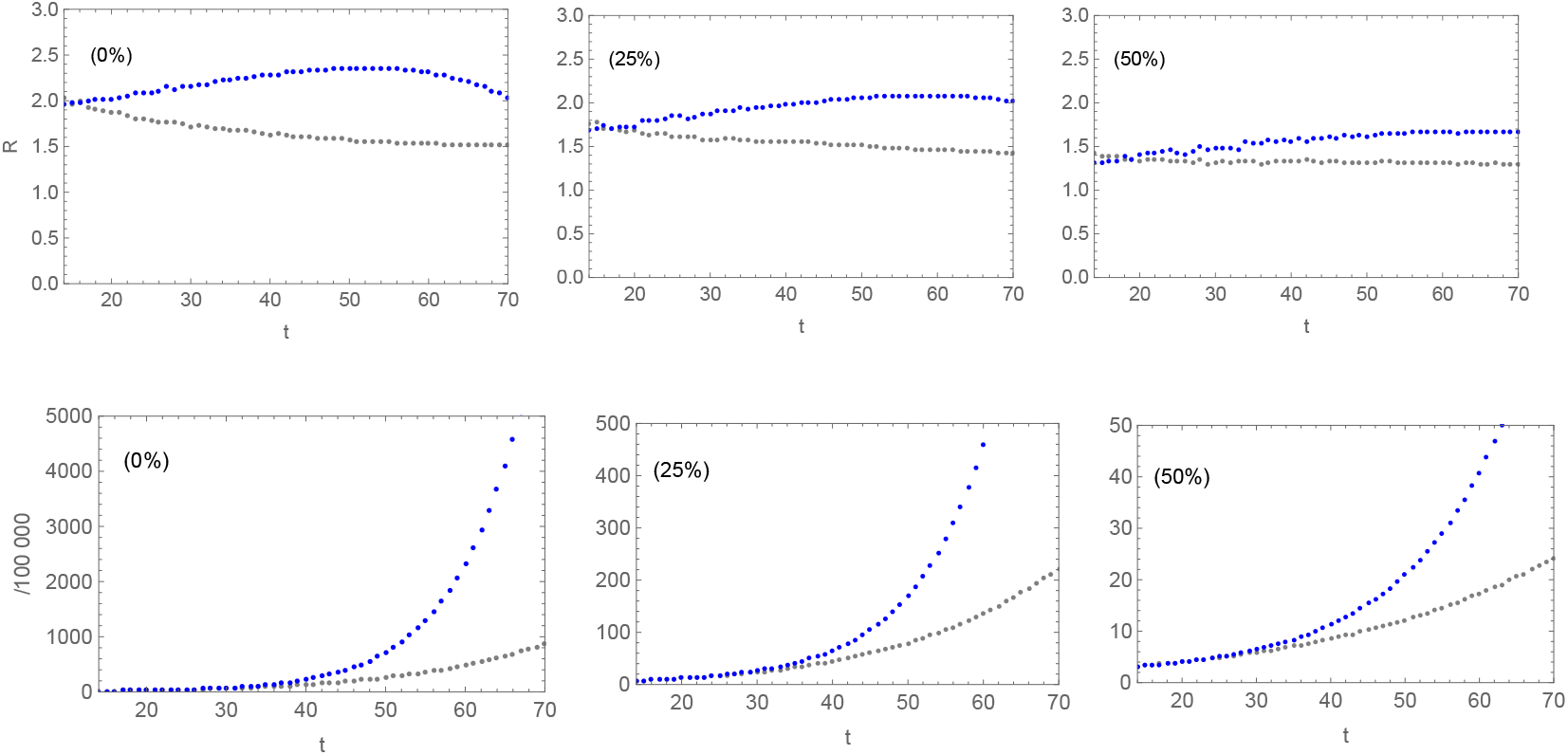
Comparison between results from the PERCOVID (gray points) and SEIR-LIKE (blue points) simulations for three different vaccination rates (0%, 25%, 50%) and for an initial incidence rate of T=2.5. The first line corresponds to the reproduction number R while the last line corresponds to the symptomatic weekly incidence rate.

**Figure 8.**
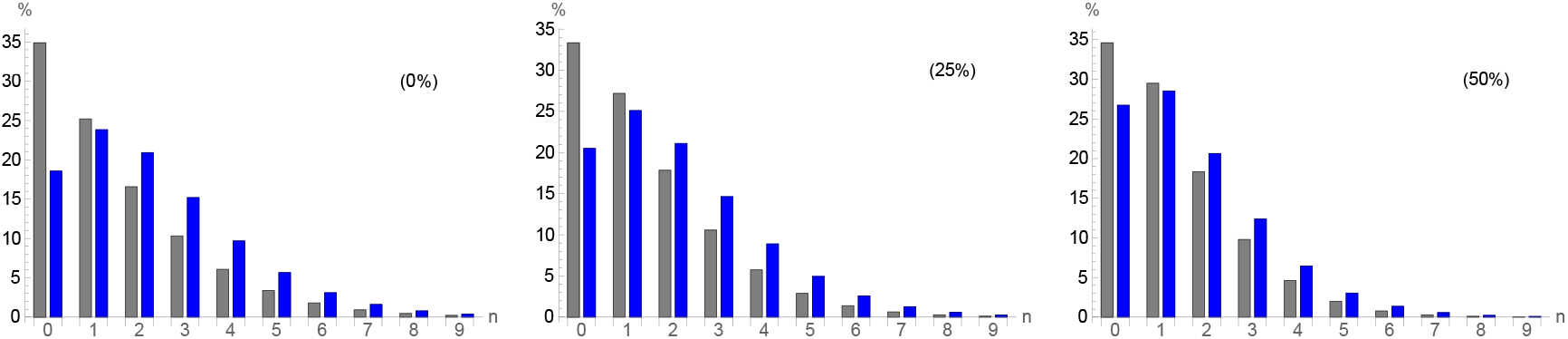
Comparison between the percentage of secondary infections. in the PERCOVID (gray bars) and SEIR-LIKE (blue bars) simulations for three different vaccination rates (0%, 25%, 50%), for an initial incidence rate of T=2.5.

- The explosion of infections found in the SEIR-LIKE simulation is also present. It is however much favored by the increase of the number of initially infected persons in the absence of vaccination, with an incidence rate of T_0_=2.5 as compared to T_0_=0.25. This leads consequently to a more rapid saturation of the reproduction number in this simulation.
- Increasing the vaccination rate reduces the population susceptible to be infected, so the explosion of infection is drastically reduced when the rate of vaccination increases. As a direct consequence, the difference in the reproduction number R in both simulations also decreases, as can be seen on figure 7.
- The percentage of secondary infections is also very similar, with an increased number of secondary infections in the SEIR-LIKE infections. These differences also reduce when increasing the number of vaccinated persons, as can be seen on figure 8.

## Discussion and conclusions

The comparison of SARS-2 propagation using a percolation and a SEIR model on four different periods of COVID-19 epidemic in France confirms the relevance but also the limits of epidemic predictions using SEIR models, especially in the early days of an epidemic. Using the flexibility and richness of the percolation model used in the PERCOVID simulation, we can manage a direct comparison between PERCOVID and SEIR models as far as the propagation pattern of the virus in each model is concerned. The PERCOVID simulation is based on a local spread of the epidemic originating from the social relationships among the population while this propagation occurs in the SEIR models all over the territory under consideration, independently of any information concerning a possible close contact between infected persons. The key lessons coming out of this comparison are the following:

i. a very large difference is found in the dynamic of the epidemic in early stages of propagation, both for the initial propagation or the spread of the delta variant, with a drastic over-estimate of the number of infected persons in the SEIR-LIKE simulation. This period is precisely the most crucial in policy making for preventing any epidemic to propagate on a large space-time scale. These features are indeed expected from the different propagation patterns of both simulations: the local and social environment of each infected person is crucial during this period where the virus propagates on a small space scale. The percolation features of PERCOVID are thus able to fully account for this behavior.
ii. When the virus is widely circulating over the territory under consideration, with an important mobility of persons together with an ambitious vaccination campaign, both simulations show similar results once the initial conditions in the SEIR-LIKE simulations are fixed by data in a typical period of two weeks before the predictions of the model.
iii. The clear separation in the PERCOVID simulation of the intrinsic properties of the virus from any social behavior of the population under study is necessary in order to access the intrinsic virulence of the virus, represented in our simulation by the intrinsic infectiousness *r*. The comparison of PERCOVID and SEIR-LIKE simulations in this study shows clearly that the intrinsic infectiousness of the virus is drastically underestimated in the SEIR-LIKE simulation, in particular in the very first stage of the propagation of the virus.

The features of PERCOVID recalled above are of particular interest as far as the propagation of respiratory track epidemics are concerned. The first stage of the underground propagation of the virus is of particular interest. The virus started propagating in the European population weeks before patients were diagnosed with severe illness, as shown in France by the cohort of healthy population CONSTANCES [13] as well as the retrospective cohort done in the Haut-Rhin department where the first major outbreak occurred [14]. In both cases the delay between the first cases and the official outbreak is more than two months. This early stage is essential in order to predict the propagation of the virus on a long-range time scale, starting from the very beginning of the propagation of the virus when it is yet unnoticed. This is in complete opposition to the extrapolation of the epidemic dynamic from data, as happens in the SEIR simulations for instance.

PERCOVID is the first model able to describe the very first steps of an epidemic without overestimating the epidemic spread and speed, therefore allowing adoption of more relevant public health policies. All these features should enable to use PERCOVID for a new insight into health policy as far as the emergence of epidemics of respiratory tracks are concerned. This possibility will be addressed in a forthcoming publication.

## Data Availability

The code PERCOVID is the property of our funding agency and cannot be made public at that time.

## Statements & Declarations

### Author Contributions

Conceptualization, L. Gerbaud and J.-F. Mathiot; methodology, V. Breton, L. Gerbaud and J.-F. Mathiot; Code programming, J.-F. Mathiot; writing original draft preparation, J.-F. Mathiot; writing, review and editing, V. Breton, L. Gerbaud and J.-F. Mathiot; visualization, J.-F. Mathiot; supervision, J.-F. Mathiot; project administration, V. Breton, L. Gerbaud and J.-F. Mathiot. All authors have read and agreed to the published version of the manuscript.

15.

## References

1. Zhang P, Feng K, Gong Y, Lee J, Lomonaco S, Zhao L. Usage of Compartmental Models in Predicting COVID-19 Outbreaks. AAPS J. 2022 Sep 2;24(5):98. Doi: 10.1208/s12248-022-00743-9. PMID: 36056223; PMCID: PMC9439263.

2. W. Kermack and A. McKendrick, 1927. A contribution to the mathematical theory of epidemics, Proc. R. Soc. A 115(772): 700–721.

3. Stauffer, D. & Aharony, A. Introduction to Percolation Theory, Crc Press (1994)

4. Sander M, Warren C P, Sokolo IM, Simon C, Koopman J. Percolation on heterogeneous networks as a model for epidemics. Math Biosci. 180, 293–305 (2002); 10.1016/s0025-5564(02)00117-7.

5. Rahimi I, Chen F, Gandomi AH. A review on COVID-19 forecasting models. Neural Comput Appl. (2023) 35(33):23671–81. 10.1007/s00521-020-05626-8

6. Salje H, et al. Estimating the burden of SARS-CoV-2 in France, Science 369, 208–211 (2020).

7. Sudhakar T, Bhansali A, Walkington J, Puelz D. The disutility of compartmental model forecasts during the COVID-19 pandemic. Front Epidemiol. 2024 Jun 20;4:1389617. Doi: 10.3389/fepid.2024.1389617. PMID: 38966521; PMCID: PMC11222405.

8. Melikechi O, Young AL, Tang T, Bowman T, Dunson D, Johndrow J. Limits of epidemic prediction using SIR models. J Math Biol. 2022 Sep 20;85(4):36. Doi: 10.1007/s00285-022-01804-5. PMID: 36125562; PMCID: PMC9487859.

9. Telle RC, Lopes H, Franco D. SARS-COV-2: SIR Model Limitations and Predictive Constraints. Symmetry 13, 676 (2021); 10.3390/sym13040676.

10. Roberts M, Andreasen V, Lloyd A, Pellis L. Nine challenges for deterministic epidemic models. Epidemics. 2015 Mar;10:49–53. Doi: 10.1016/j.epidem.2014.09.006.

11. Mathiot J-F, Gerbaud L, Breton V. Highlighting the impact of social relationships on the propagation of respiratory viruses using percolation theory; Scientific Reports 11, 24326 (2021).

12. Cori A, Ferguson NM, Fraser Ch, Cauchemez S. A New Framework and Software to Estimate Time-Varying Reproduction Numbers During Epidemics, American Journal of Epidemiology, Volume 178, Issue 9, 1 November 2013, Pages 1505–1512, 10.1093/aje/kwt13.

13. Carrat F, Figoni J, Henny J, Desenclos JC et al. Evidence of early circulation of SARS-CoV-2 in France: findings from the population-based “CONSTANCES” cohort. Eur J Epidemiol 2021;36(2):219–222; 10.1007/s10654-020-00716-2.

14. Gerbaud L, Guiguet-Auclair C, Breysse F et al. Hospital and Population-Based Evidence for COVID-19 Early Circulation in the East of France. Int J Environ Res Public Health 2020, 30;17(19):7175; 10.3390/ijerph17197175.

